## Supplement for "Exploring the Spatial Distribution of Persistent SARS-CoV-2 Mutations - Leveraging mobility data for targeted sampling"

### Table of Contents


1. Supplementary Files
    1.1. Supplementary File 1: “microreact”-file summarizing all data presented in the microreact-project.
    1.2. Supplementary File 2: “xz”-packed fasta-file, containing all SARS-CoV-2 Alpha lineage genomes used in the Nextstrain analysis
    1.3. Supplementary File 3: Metadata tsv-file containing the information for all SARS-CoV-2 Alpha lineage genomes used in the Nextstrain analysis.
    1.4. Supplementary File 4: “yaml”-file containing the build-instructions for the Nextstrain analysis.
    1.5. Supplementary File 5: “xz”-packed fasta-file with the resulting, subsampled genomes from the Nextstrain analysis.
    1.6. Supplementary File 6: Nextstrain “tree.nwk”-file used to visualize the phylogenetic time tree.
    1.7. Supplementary File 7: Metadata tsv-file for the Alpha-lineage genomes contained in the phylogenetic time tree.
2. Supplementary Tables
    2.1. Supplementary Table S1: Total number of reported SARS-CoV-2 samples and number of reported Alpha lineage samples for each German federal state per month.
    2.2. Supplementary Table S2: Total number of reported SARS-CoV-2 samples and number of reported Alpha lineage samples for each Thuringian district per month.
    2.3. Supplementary Table S3: Accumulated sample count for each Thuringian district per Alpha subcluster and per month.
    2.4. Supplementary Table S4: Total number of incoming trips and numbers of trips coming from all cluster-affiliated districts to each Thuringian district per Alpha subcluster and per month.
    2.5. Supplementary Table S5: Overview of all Thuringian samples collected for the mobility-guided pilot experiment between the 5th of October and 25th of November 2022.
    2.6. Supplementary Table S6: Mobility data and sampling counts of communities sampled for the mobility-guided pilot experiment between the 5th of October and 25th of November 2022.
3. Supplementary Figures
    3.1. Supplementary Figure S1: Total population (A) and population density per km^2 (B) for each Thuringian district as stated for the 31st of December 2020.
    3.2. Supplementary Figure S2: Total number of all sequenced SARS-CoV-2 samples (purple) and the proportion of the Alpha lineage for all sequenced samples (yellow-red) for each state of Germany and each district of Thuringia throughout the whole observation period.
    3.3. Phylogenetic time tree construction
    3.4. Supplementary Figure S3: Phylogenetic time tree of the Alpha lineage.
    3.5. Supplementary Figure S4: Accumulated number of sequenced samples for each Alpha lineage subcluster per district and per month.
    3.6. Supplementary Figure S5: Combined visualization of each district's “inbound mobility” from other districts (color intensity) and the occurrence of a subcluster sample (red = sample found, blue = no sample found) per subcluster.
4. References

### 1. Supplementary Files

All supplementary files are provided as part of an OSF project under the following link: https://osf.io/n5qj6/ (DOI: 10.17605/OSF.IO/N5QJ6).

**Supplementary File 1: "microreact"-file summarizing all data presented in the microreact-project.** The project can be found at https://microreact.org/project/ftR2GfjF6iXtSwbmN4ARTx-thuringianalpha-linclusters#76ir-complete-overview.

**Supplementary File 2: "xz"-packed fasta-file, containing all SARS-CoV-2 Alpha lineage genomes used in the Nextstrain analysis**

**Supplementary File 3: Metadata tsv-file containing the information for all SARS-CoV-2 Alpha lineage genomes used in the Nextstrain analysis.**

**Supplementary File 4: "yaml"-file containing the build-instructions for the Nextstrain analysis.**

**Supplementary File 5: "xz"-packed fasta-file with the resulting, subsampled genomes from the Nextstrain analysis.**

**Supplementary File 6: Nextstrain "tree.nwk"-file used to visualize the phylogenetic time tree.** Contains 64,131 German (non-Thuringian) and 6,298 Thuringian Alpha-lineage genomes.

**Supplementary File 7: Metadata tsv-file for the Alpha-lineage genomes contained in the phylogenetic time tree.**

### 2. Supplementary Tables

All supplementary tables are provided as "tsv"-files in an OSF project under the following link: https://osf.io/n5qj6/ (DOI: 10.17605/OSF.IO/N5QJ6).

**Supplementary Table S1: Total number of reported SARS-CoV-2 samples and number of reported Alpha lineage samples for each German federal state per month**. The respective proportions of Alpha and non-Alpha samples are calculated.

**Supplementary Table S2: Total number of reported SARS-CoV-2 samples and number of reported Alpha lineage samples for each Thuringian district per month.** The respective proportions of Alpha and non-Alpha samples are calculated.

**Supplementary Table S3: Accumulated sample count for each Thuringian district per Alpha subcluster and per month.**

**Supplementary Table S4: Total number of incoming trips and numbers of trips coming from all cluster-affiliated districts to each Thuringian district per Alpha subcluster and per month.** The proportion of trips coming from cluster-affiliated districts among the total incoming trips is calculated.

**Supplementary Table S5: Overview of all Thuringian samples collected for the mobility-guided pilot experiment between the 5th of October and 25th of November 2022.**

**Supplementary Table S6: Overview of the communities sampled for the mobility-guided pilot experiment between the 5th of October and 25th of November 2022.** For each community, the incoming one-way trips from the community of BQ.1.1 origin in Thuringia based on the October 2020 and June 2021 datasets are provided. Additionally, the number of guided and randomized samples per community and an indication of found BQ.1.1 samples are provided.

### 3. Supplementary Figures

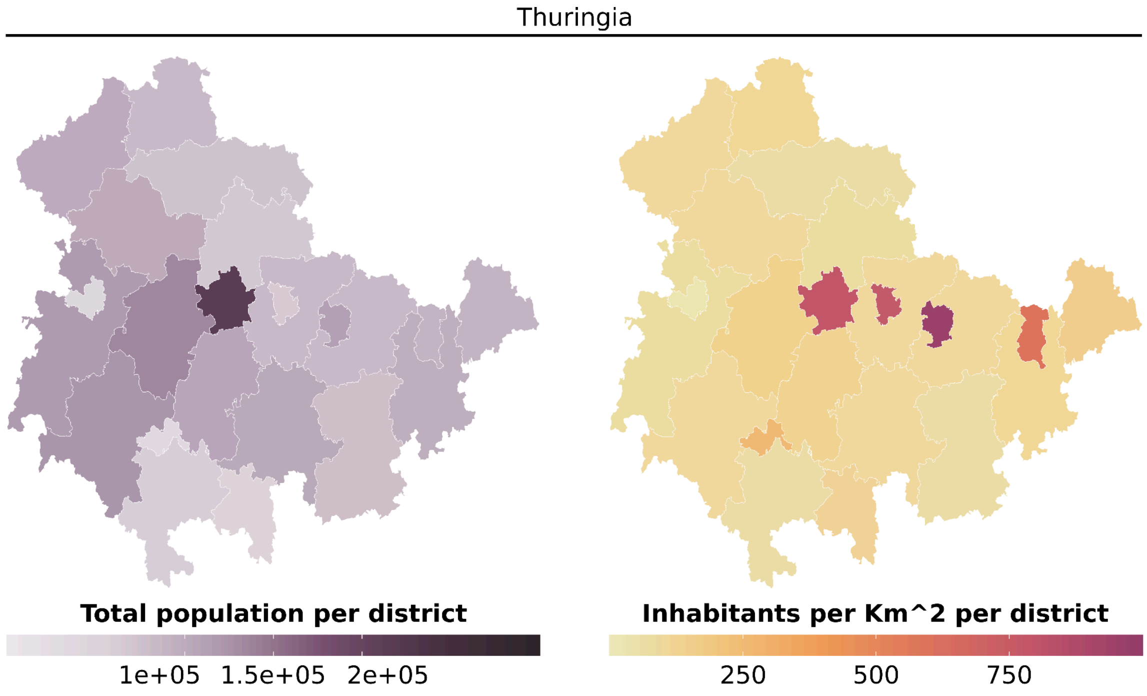


***Supplementary Figure S1: Total population (A) and population density per km^2 (B) for each Thuringian district as stated for the 31st of December 2020.***

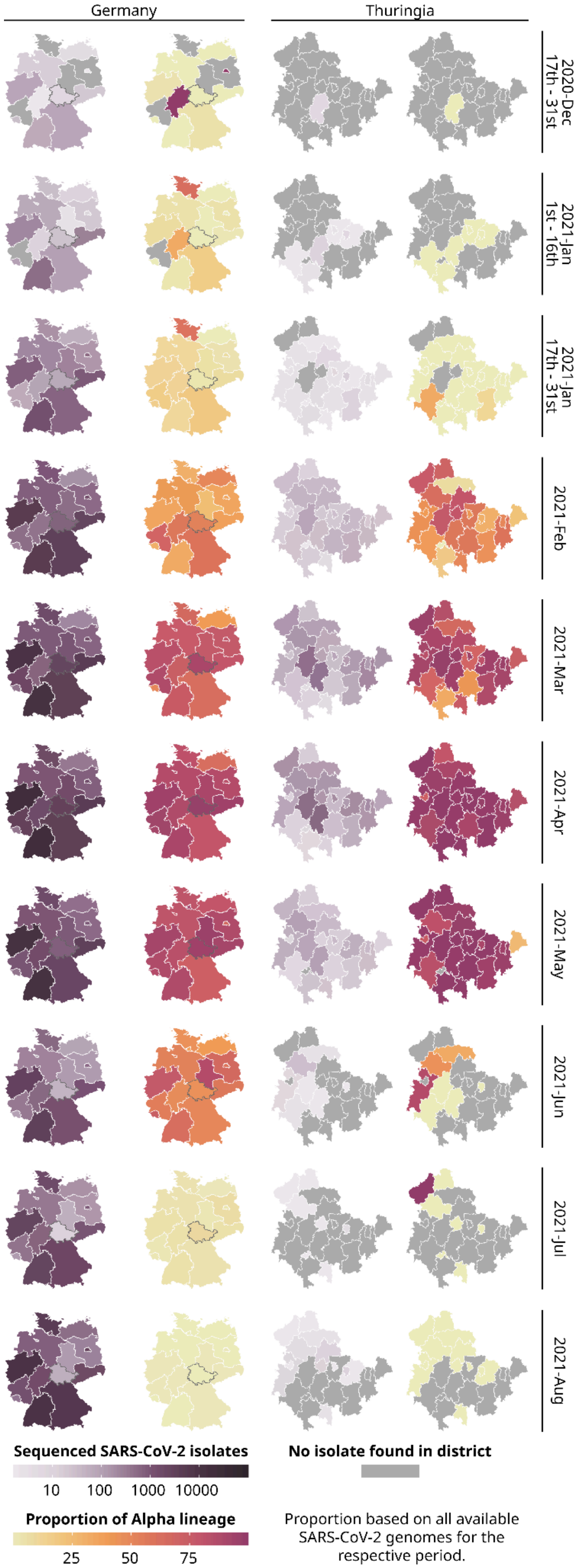
Germany
Thuringia
2020-Dec 17th - 31st
2021-Jan 1st - 16th
2021-Jan 17th - 31st
2021-Feb
2021-Mar
2021-Apr
2021-May
2021-Jun
2021-Jul
2021-Aug
Sequenced SARS-CoV-2 isolates
10 100 1000 10000
No isolate found in district
Proportion of Alpha lineage
25 50 75
Proportion based on all available SARS-CoV-2 genomes for the respective period.

***Supplementary Figure S2: Total number of all sequenced SARS-CoV-2 samples (purple) and the proportion of the Alpha lineage for all sequenced samples (yellow-red) for each state of Germany and each district of Thuringia throughout the whole observation period.*** *289,487 publicly available German SARS-CoV-2 genomes and their metadata were used for the general German maps excluding data from Thuringia. For Thuringia, we always used 7,394 genomes and their metadata from our database for both the German and the Thuringian maps. Please note that for all states except Thuringia, we used the postal code of the sending laboratory as a proxy for the geographic location of a sample. Thuringia is highlighted by a grey border on the maps of Germany.*

#### Phylogenetic time tree construction

7,262 sequenced Alpha lineage genomes with less than 12,000 ambiguous bases were exported with the respective data from our local database. These were filtered to remove 609 non-Thuringian isolates, four duplicates, and 127 entries missing location information or isolation date. After filtering, the Thuringian dataset contained 6,522 isolates.

From the 1,091,655 public German genomes provided by the Robert Koch Institute, 146,347 Alpha lineage isolates were extracted. For these, more specific location data (country, state, district, city, latitude, longitude) were added based on the sending laboratory's postal code. Subsequently, 9,600 Thuringian isolates were removed. The resulting 136,747 genomes were then analyzed with poreCov (version 1.7.2) to determine mutations/deletions and confirm the lineages.[1] The results were uploaded to a separate collection in our local database. poreCov identified a further 108 non-Alpha isolates, which were subsequently removed from the dataset. During the generation of a test phylogenetic time tree (see below), "Nextstrain" excluded 540 isolates due to quality issues.[2] After their removal, the final German Alpha dataset comprised 136,099 entries.

The prepared Thuringian and German Alpha lineage datasets were combined to build a phylogenetic time tree using the program "Nextstrain" (version 5.0.0).[2] Following the program's instructions, two genomes of the original Wuhan lineage were added to the combined fasta-file, while their respective data were added to the combined data file (omitting this step would lead to a crash in the used Nextstrain version). It was ensured that the data file contained the necessary columns given in the manual. Further, in our version, the data file needed to contain at least 12 columns, or Nextstrain would have an error. The resulting fasta and metadata file for the nextstrain analysis are provided as Supplementary Files 2 and 3, respectively.

A custom "build.yaml"-file was used for the nextstrain analysis, which is provided as Supplementary File 4:

```
inputs:
  - name: alpha-lin_phylotree
     metadata: data/alpha-lin_phylotree_metadata.tsv
     sequences: data/alpha-lin_phylotree.fasta
```

```
builds:
  thuringia:
      subsampling_scheme: custom-division
      region: Europe
      country: Germany
      division: Thuringia
subsampling:
  custom-division:
      focal:
      group_by: "year month"
      seq_per_group: 10000000
      query: --query "(country == '{country}') & (division ==
'{division}')"

  related:
      group_by: "country year month"
      seq_per_group: 16000
      exclude: "--exclude-where 'division={division}'"
      priorities:
      type: "proximity"
      focus: "focal"
files:
                                                    auspice_config:
"my_profiles/alpha-lin_analysis/my_auspice_config.json"
  description: "my_profiles/alpha-lin_analysis/my_description.md"
```

The analysis started with the command:
"`nextstrain build . --cores 32 --profile my_profiles/path/to/build/dir`".

The resulting phylogenetic time tree comprises 64,131 German (non-Thuringian) and 6,298 Thuringian Alpha-lineage genomes. Additional 2,160 German (non-Thuringian) and 224 Thuringian genomes are included in the metadata but not visualized in the tree and, therefore, not considered for the following analysis. The resulting tree is shown in Supplementary Figure S3 below. The subsampled genome-, tree-, and metadata file are provided as Supplementary Files 5, 6, and 7.

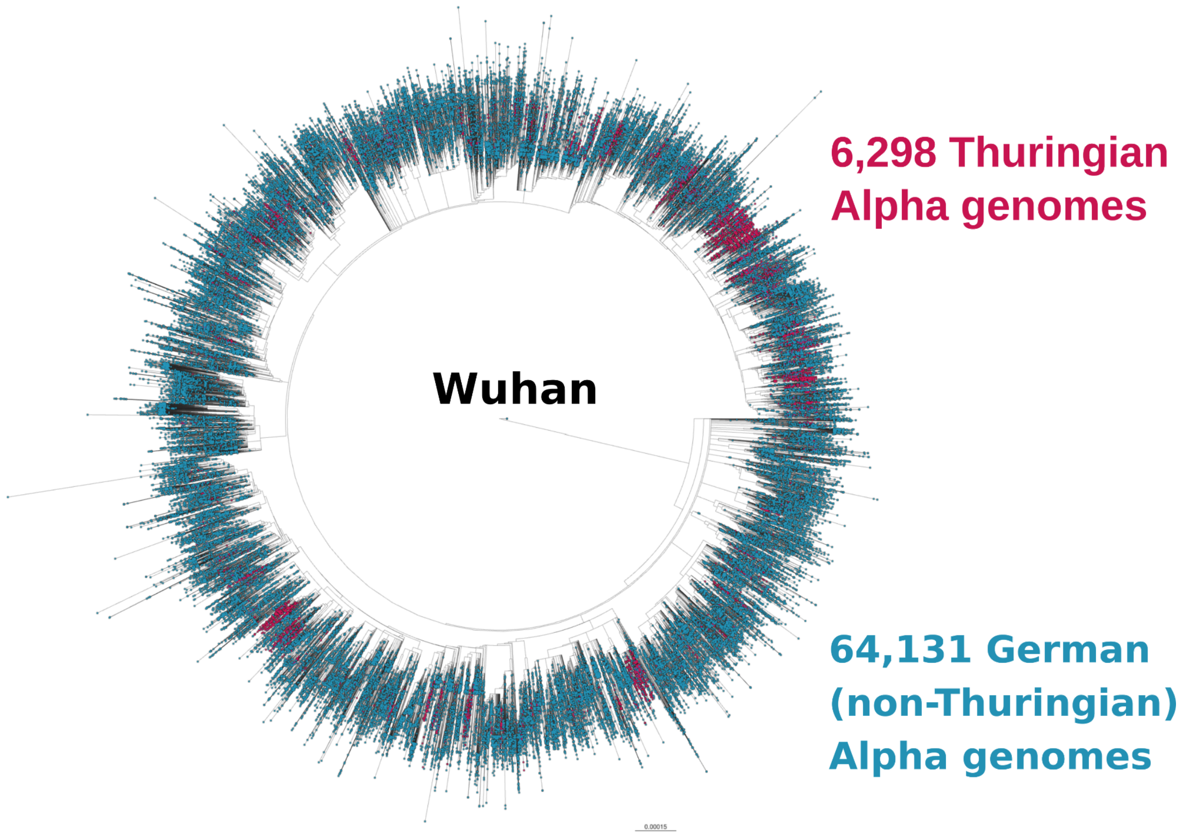


***Supplementary Figure S3: Phylogenetic time tree of the Alpha lineage.*** *The tree includes 64,131 German (non-Thuringian; blue) and 6,298 Thuringian (red) Alpha genomes. Two genomes of the original Wuhan lineage are included as origin.*

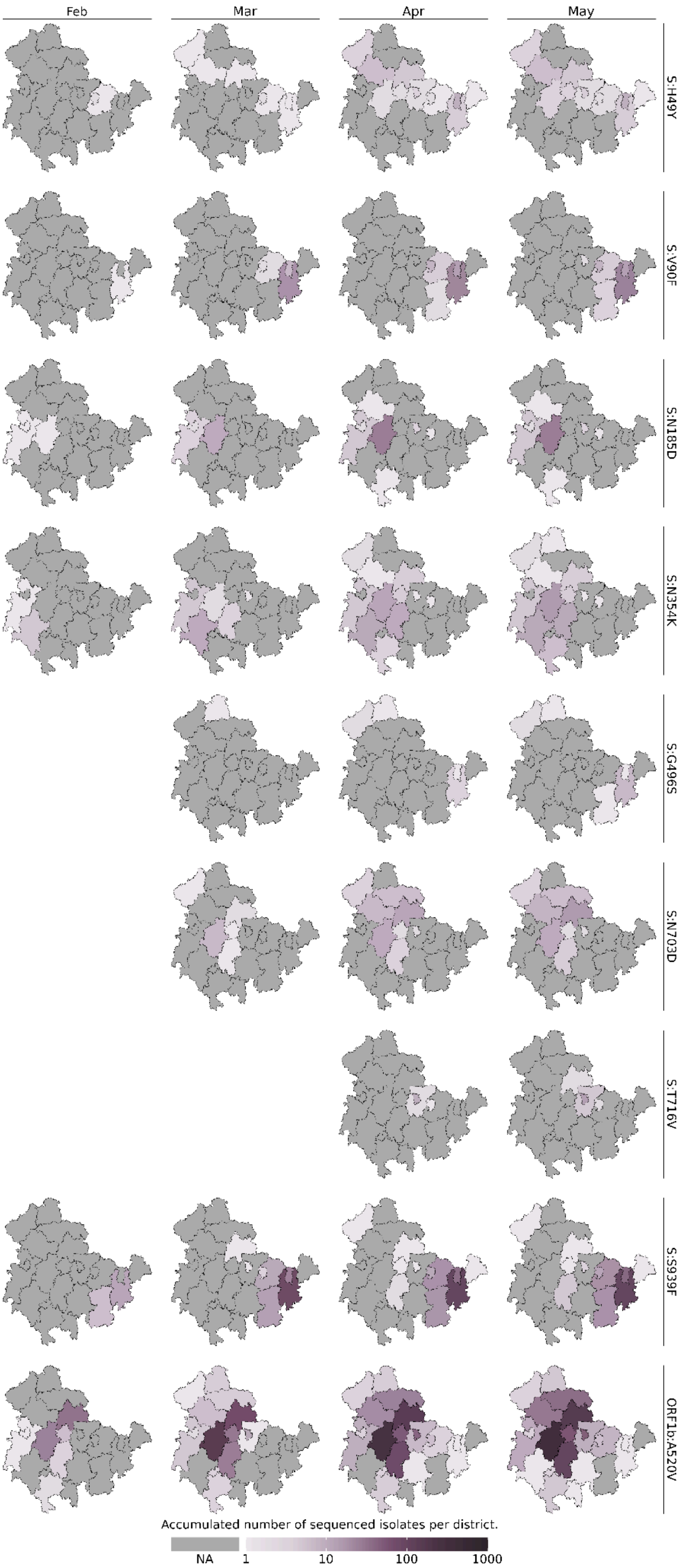


***Supplementary Figure S4: Accumulated number of sequenced samples for each Alpha lineage subcluster per district and per month.***

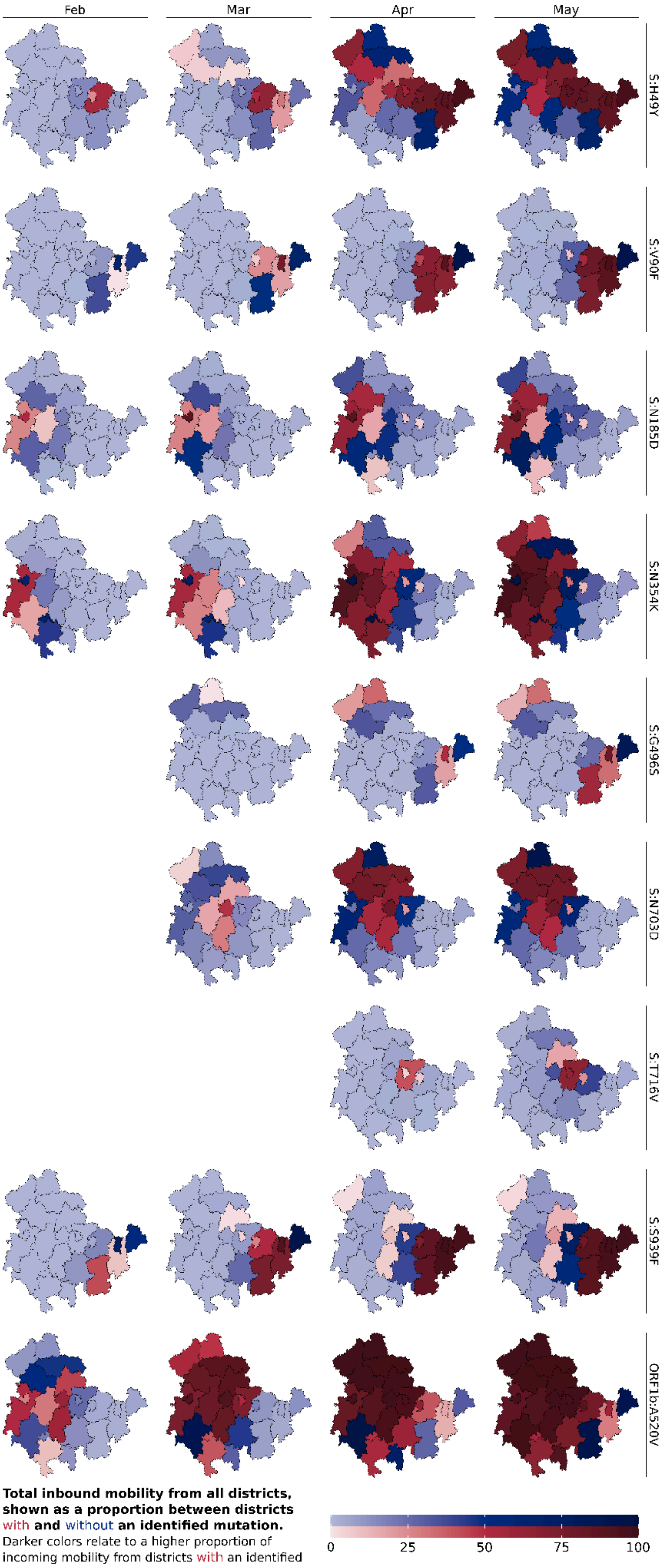


**Total inbound mobility from all districts, shown as a proportion between districts with and without an identified mutation.**
Darker colors relate to a higher proportion of incoming mobility from districts with an identified mutation

***Supplementary Figure S5: Combined visualization of each district's "inbound mobility" from other districts (color intensity) and the occurrence of a subcluster sample (red = sample found, blue = no sample found) per subcluster.*** *The inbound mobility of each district (color intensity) is shown as a proportion of incoming mobility from other districts with or without an identified sample. The darker the color (red and blue) of a district, the higher the proportion of inbound mobility from other districts with an identified subcluster sample (red districts). The light blue color describes that most of the inbound mobility of a district comes from other districts without an identified subcluster sample (blue districts).*
